# Evaluation of a structured screening assessment to detect patients with isolated REM Sleep Behavior Disorder

**DOI:** 10.1101/2022.10.23.22281409

**Authors:** Aline Seger, Anja Ophey, Wiebke Heitzmann, Christopher E. J. Doppler, Marie-Sophie Lindner, Corinna Brune, Johanna Kickartz, Haidar S. Dafsari, Wolfgang H. Oertel, Gereon R. Fink, Stefanie T. Jost, Michael Sommerauer

**Affiliations:** University of Cologne, Faculty of Medicine and University Hospital Cologne, Department of Neurology, Cologne, Germany; Cognitive Neuroscience, Institute of Neuroscience and Medicine (INM-3), Research Centre Jülich, Jülich, Germany; University of Cologne, Faculty of Medicine and University Hospital Cologne, Medical Psychology, Neuropsychology and Gender Studies & Center for Neuropsychological Diagnostics and Interventions (CeNDI), Cologne, Germany; Department of Neurology, Philipps-University Marburg, Marburg, Germany

**Keywords:** RBD, Parkinson’s disease, prognostic model, questionnaire

## Abstract

**Background:** Isolated rapid eye movement (REM) sleep behavior disorder (iRBD) cohorts have provided novel insights in the earliest neurodegenerative processes in α-synucleinopathies. Even though polysomnography remains the gold standard for diagnosis, an accurate questionnaire-based algorithm to identify eligible subjects could facilitate efficient recruitment in research.

**Objectives:** This study aimed to optimize the identification of subjects with iRBD from the general population.

**Methods:** Between June 2020 and July 2021, we placed newspaper advertisements including the single-question screen for RBD (RBD1Q). Participants’ evaluations included a structured telephone screening consisting of the RBD screening questionnaire (RBDSQ) and additional sleep-related questionnaires. We examined anamnestic information predicting polysomnography-proven iRBD using logistic regressions and receiver operating characteristic curves.

**Results:** 543 participants answered the advertisements and 185 subjects fulfilling in- and exclusion criteria were screened. Of these, 124 received polysomnography after expert selection and 78 (62.9%) were diagnosed with iRBD. Selected items of the RBDSQ, the Pittsburgh Sleep Quality Index, the STOP-Bang questionnaire, and age predicted iRBD with high accuracy in a multiple logistic regression model (area under the curve >80%). Comparing the algorithm to the sleep expert decision, 77 instead of 124 polysomnographies (62.1%) would have been carried out, while 63 (80.8%) of iRBD patients would have been identified. 32 of 46 (69.6%) unnecessary polysomnography examinations could have been avoided.

**Conclusions:** Our proposed algorithm displayed high diagnostic accuracy for polysomnography-proven iRBD in a cost-effective manner and may be a convenient tool for application in research and clinical settings. External validation sets are warranted to prove its reliability.

## Introduction

Isolated rapid eye movement (REM) sleep behavior disorder (iRBD) is considered as a highly specific marker of incipient α-synucleinopathy.^1–3^ Longitudinal studies indicate that more than 80% of iRBD subjects eventually develop Parkinson’s disease (PD), dementia with Lewy-bodies (DLB) or Multiple system atrophy (MSA).^4^ iRBD cohorts have provided novel insights in the early neurodegenerative processes in α-synucleinopathies and have already played a major role in understanding corresponding neuropathological mechanisms.^5–7^ The reliable identification of iRBD subjects in the general population is essential to study the fundamental sequences of neurodegeneration in the time course of early α-synucleinopathies towards conversion to manifest motor disease or dementia.

The gold standard of RBD diagnosis is video-polysomnography (PSG). However, execution of PSG is labor-intensive and costly. Hence, a well-guided selection of subjects with a high *a priori* probability of having iRBD is required. A widely used and previously validated screening tool to evaluate RBD symptoms is the RBD screening questionnaire (RBDSQ).^8^ However, recent studies questioned the diagnostic accuracy of the RBDSQ in the identification of RBD in unselected cohorts and in *de novo* PD patients.^9,10^

Due to the lack of a precise screening tool, the selection of subjects to PSG during the screening process is typically made by a sleep expert as an important first step. However, such subjective judgments are mostly elusive, and typically multiple PSGs are needed to identify a single iRBD subject. Additionally, such subjective decision-making processes are naturally difficult to be generalized and to be transferred to other research-centers, e.g., with limited sleep-medicine resources and expertise. Therefore, an accurate and easy to apply algorithm to identify subjects with a high probability to have iRBD remains quested for efficient recruitment of iRBD cohorts in research. This might even become more compelling in the near future, when disease-modifying drugs may be available.^11^ A more targeted use of PSG has the potential to reduce costs, save resources, and eventually facilitate iRBD cohort recruitment.

With the present work, we aimed to optimize the identification of subjects with iRBD from the general population with a novel screening algorithm combining multiple already available questionnaires, covering aspects relevant for the differential diagnosis of iRBD by identifying the most relevant items of those questionnaires. Thereby, we wanted to facilitate the selection of suited patients for PSG and reduce its demand. We conducted a structured questionnaire-based screening assessment and evaluated the accuracy of items in predicting iRBD. Following, we developed a two-step algorithm and made it accessible for researchers and clinicians.

## Material and methods

### Study design and ethical approval

Subjects were recruited between June 2020 and July 2021 to establish a local iRBD cohort at the Department of Neurology of the University Hospital Cologne. The study protocol was approved by the local ethics committee (vote number 19-1408) and subjects provided written informed consent according to the Declaration of Helsinki.

### Screening procedure

Subjects were recruited via graphical advertisements showing stereotypical signs of RBD (elderly man with dream-enacting behavior and violent dream content, example is given in the supplementary material) and the German version of the single-question screen for RBD (RBD1Q; “Did you notice or did someone else notice that you do act out your dreams?”) in local newspapers.^12^ Subjects, who could affirm to the RBD1Q, were encouraged to contact our research group. Prior to further screening, all subjects were informed about the scope of the study and the potential risk of being diagnosed with iRBD as a potential α-synucleinopathy to allow for early dropout in case the subject did not want to know about its individual risk of having a neurodegenerative disease.

In- and exclusion criteria for full screening were as following: Inclusion, RBD1Q answered with “yes”; exclusion, known neurological disorder (i.e., PD, epilepsy with seizures during the night, and narcolepsy), age < 35 years or > 80 years, early age of symptom onset (< 35 years), alcohol or drug abuse, and having a pacemaker.

If not meeting any exclusion criteria, subjects were asked to provide further demographic data and medical history. Additionally, the following validated self-rating scales and questionnaires were assessed to cover a broad range of sleep disturbances: (i) the full *RBDSQ*, (ii) the *Pittsburgh Sleep Quality Index (PSQI)*^*13*^ providing seven component scores including subjective sleep quality, sleep latency, sleep duration, habitual sleep efficiency, sleep disturbances, use of sleeping medication, and daytime dysfunction as well as a global sleep quality score, (iii) the *STOP-Bang*^*14*^ questionnaire, a screening tool for obstructive sleep apnea, (iv) the *Epworth Sleepiness Scale*^*15*^ *(ESS)* to screen for daytime sleepiness, and (v) the *Regensburg Insomnia Scale*^*16*^ *(RIS)* to assess various aspects of insomnia. To screen for presence of symptoms of the Restless Legs Syndrome (RLS), (vi) a 10-item self-test published by the German RLS Society (https://www.restless-legs.org/restless-legs/syndrom/selbsttest/) was applied covering the diagnostic criteria for RLS.^17^

All information gained from the structured telephone screening were evaluated by a neurologist and board-certified sleep-expert with long standing experience in iRBD research (MS).^18^ Based on his evaluation, selected subjects were invited to PSG to confirm the diagnosis of iRBD according to consensus criteria from the International Classification of Sleep Disorders III.^19^

### Polysomnography

All PSGs were undertaken at home or in a hotel room. We used a mobile SOMNOscreen™ plus device for overnight video-PSG including 10 EEG recordings (according to the international 10/20 system: F3, F4, C3, C4, O1, O2, A1, A2, Fpz as grounding, and Cz as reference), electrooculography, surface EMG of the mental, the tibialis anterior, and flexor digitorum superficialis muscles, electrocardiography, nasal pressure and flow monitoring, thoracic and abdominal respiratory effort belts, finger pulse oximetry, and synchronized audio-visual recording. Before lights off, electrode impedances were checked to be lower than 10kΩ.

Visual PSG scoring was performed on 30-second epochs including sleep efficiency, total sleep time, absolute amount of stage 1 (N1), stage 2 (N2), stage 3 (N3), and REM sleep, apnea-hypopnea index (AHI, number of apnea plus hypopnea events per hour of sleep) and periodic limb movement index (PLMI, number of periodic leg movements per hour of sleep) according to the American Academy of Sleep Medicine (AASM) Manual for the Scoring of Sleep and Associated Events Version 2.6.^20^

### Statistical analysis

Statistical analyses were performed using IBM SPSS Statistics 28 and RStudio (R version 4.2.0) with the purpose to reassess the decision-making process of the sleep expert to select subjects for PSG **(step 1)** and to identify subjects with iRBD at PSG **(step 2)**. Subjects with completed screening were divided into three subgroups: (1) participants who underwent a completed telephone screening but were not invited to PSG due to the expert’s evaluation, (2) participants who received PSG but were not diagnosed with iRBD, and (3) participants who received PSG and were eventually diagnosed with iRBD (Table 1).

**Table 1.** Clinical and demographic characteristics of subgroups.

| Characteristics | All subjects<br>(n=185) | Subgroup 1:<br>No PSG<br>(n=61) | Selected for PSG by sleep expert |  | *p*<sup>†</sup> |
| --- | --- | --- | --- | --- | --- |
|  |  |  | Subgroup 2:<br>PSG, no iRBD<br>(n=46) | Subgroup 3:<br>PSG and iRBD<br>(n=78) |  |
| Age | 63.5 ± 9.8 | 62.5 ± 10.6 | 60.5 ± 12.1 | 66.1 ± 6.7 | **0.024<sup>b</sup>** |
| Sex, female (%) | 37 (20) | 13 (21.3) | 13 (28.3) | 11 (14.1) | 0.155 |
| BMI | 26.2 ± 3.8 | 27.4 ± 4.7 | 26.2 ± 3.1 | 25.3 ± 3.3 | **0.011<sup>a</sup>** |
| RBDSQ total score | 8.8 ± 2.3 | 7.7 ± 2.5 | 8.7 ± 2.1 | 9.7 ± 1.9 | **<0.001<sup>a, b</sup>** |
| STOP-Bang total score | 3.5 ± 1.5 | 4.0 ± 1.8 | 3.2 ± 1.4 | 3.2 ± 1.4 | **0.021<sup>a</sup>** |
| ESS total score | 7.8 ± 3.7 | 8.2 ± 3.8 | 8.0 ± 3.4 | 7.5 ± 3.8 | 0.439 |
| RLS total score | 2.5 ± 2.4 | 3.2 ± 2.5 | 2.7 ± 2.5 | 1.8 ± 2.0 | **<0.001<sup>a, b</sup>** |
| PSQI total score | 7.6 ± 4.7 | 9.0 ± 5.4 | 8.1 ± 4.7 | 6.3 ± 3.7 | **0.004<sup>a, b</sup>** |
| Sleep quality | 1.3 ± 0.7 | 1.5 ± 0.7 | 1.6 ± 0.8 | 1.1 ± 0.6 | **0.001<sup>b</sup>** |
| Sleep latency | 1.0 ± 0.8 | 1.1 ± 1.1 | 0.9 ± 0.7 | 0.9 ± 0.7 | 0.969 |
| Sleep duration | 0.9 ± 0.9 | 1.1 ± 1.0 | 1.1 ± 0.9 | 0.6 ± 0.7 | **0.011<sup>b</sup>** |
| Sleep efficiency | 0.8 ± 1.0 | 1.0 ± 1.1 | 0.9 ± 1.1 | 0.6 ± 0.9 | 0.055 |
| Sleep disturbances | 1.2 ± 0.6 | 1.3 ± 0.6 | 1.2 ± 0.6 | 1.0 ± 0.4 | **0.005<sup>a, b</sup>** |
| Sleep medication | 0.2 ± 0.7 | 0.3 ± 0.8 | 0.3 ± 0.8 | 0.2 ± 0.7 | 0.191 |
| Daytime dysfunction | 0.8 ± 0.8 | 1.1 ± 0.8 | 0.6 ± 0.8 | 0.6 ± 0.7 | **<0.001<sup>a</sup>** |
| RIS total score | 8.8 ± 5.8 | 10.0 ± 6.1 | 10.0 ± 6.3 | 6.8 ± 4.7 | **<0.001<sup>a, b</sup>** |

The first subgroup did not receive PSG due to expert evaluation. The second subgroup received PSG but had no iRBD and the third subgroup - the final iRBD-cohort - had iRBD confirmed by PSG. Significant results are highlighted in bold font. Data presented are mean ± standard deviation unless indicated otherwise.
<sup>†</sup> Kruskal-Wallis tests for continuous variables and Chi-square-test for categorical data (sex) between characteristics of subgroups 1, 2 and 3. <sup>a</sup> Significant differences using Mann-Whitney *U* tests for pairwise comparisons between subgroup 1 vs. subgroups 2+3. <sup>b</sup> Significant differences using Mann-Whitney *U* tests for pairwise comparisons between subgroup 2 vs. subgroup 3.
Abbreviations: BMI, Body Mass Index; ESS, Epworth Sleepiness Scale; iRBD, isolated REM Sleep Behavior Disorder; RLS, Restless Legs Study screening questionnaire; PSG, Polysomnography; PSQI, Pittsburgh Sleep Quality Index; RBDSQ, REM Sleep Behavior Disorder Symptom Questionnaire; RIS, Regensburg Insomnia Scale; STOP-Bang, screening tool for obstructive sleep apnea.

The assumption of normality of data was inspected with Shapiro-Wilk tests. We analyzed differences of characteristics between the three subgroups using Chi-square-test for categorical data and Kruskal-Wallis tests for continuous variables. Post hoc Mann-Whitney *U* tests and *t* tests, respectively, were employed for pairwise comparisons of subgroups.

To not give too much emphasis to single items of a questionnaire, we decided to use sum scores of the questionnaires in final analysis: We first evaluated the most important items of the RBDSQ and then computed a sum score of the resulting items (see section *Results*), similarly to a previous publication.^10^ Because of the predominance of male sex in iRBD and PD, we excluded question 8 (gender) of the STOP-Bang. As the first two questions of the RIS were already included in the PSQI, we excluded these questions and built the sum of questions 3 to 10. Furthermore, we included only the component scores of the PSQI for final analysis.

We employed exploratory logistic regression analyses in a two-step procedure in order to reflect the two-stage process of detecting subjects with iRBD in the best possible way:

**(Step 1)** First, we focused on the expert rating to identify the most important variables of the selection process for receiving a PSG. In this step, all subjects were included who fulfilled in- and exclusion criteria and completed the telephone screening assessments. A dichotomous variable with “no PSG” (subgroup 1) vs. “PSG” (subgroups 2 and 3) was used as the criterion variable. Variables with *p-*values < 0.1 in simple logistic regression analyses were considered for entry into the multiple regression analysis. The final model was ascertained based on the results of multiple logistic regression by using backward stepwise elimination. Based on the predicted probabilities for the criterion variable extracted from the model, the predictive ability of the resulting model was assessed by means of a receiver-operator characteristics curve (ROC) and quantified by the C-statistic also known as the area under the curve (AUC).^21^ An AUC of 0.5 corresponds to a predictive ability equal to that of chance, whereas an AUC of 1.0 represents perfect predictive ability.

**(Step 2)** In the second step, we only included patients in the analysis who received PSG (subgroups 2 and 3) to identify the most relevant anamnestic predictors for final PSG-proven diagnosis of iRBD (after expert selection to PSG). Presence of PSG-confirmed iRBD was used as the criterion variable (“iRBD” vs. “no iRBD”) and similarly to step 1, data from the structured telephone interview were set as candidate predictors. Again, a selection process based on simple logistic regression analyses was followed by multiple logistic regression and ROC analyses.

The performance of the resulting step 1 and step 2 algorithm (step 1 for selecting participants for PSG and step 2 for predicting iRBD in subjects undergoing PSG) were compared to the prognostic accuracy of the established RBDSQ total score as a screening tool for iRBD by comparing the AUC using the *roc*.*test* function as implemented in the R (https://www.r-project.org) package *pROC* using the DeLong method.^22^

Furthermore, we applied the results of the final two-step logistic regression model to all completely screened subjects to identify potential iRBD patients by the developed algorithm who were rejected to PSG by expert ratings. These subjects were invited to PSG to additionally assess the algorithm’s performance.

### Data availability

The data included in this study are available on reasonable request to the corresponding author. The data are not publicly available due to the inclusion of information that could compromise the privacy of the participants.

## Results

### Participants’ characteristics and differences between subgroups

Results of subject selection are summarized in figure 1. 543 subjects responded to the advertisements, 329 were excluded as they did not fulfill in- and exclusion criteria, 29 were excluded from statistical analysis due to insufficient data. Eventually, 185 subjects (37 female) were eligible for further analysis. Expert evaluations selected 124 of these subjects to PSG and 78 were diagnosed with iRBD, implying a specificity of 62.9% of the expert rater to detect iRBD upon screening. Clinical and demographic characteristics of these subgroups are shown in table 1. Significant differences between the subgroups were found in all evaluated rating scales except for the ESS.

**Figure 1.**
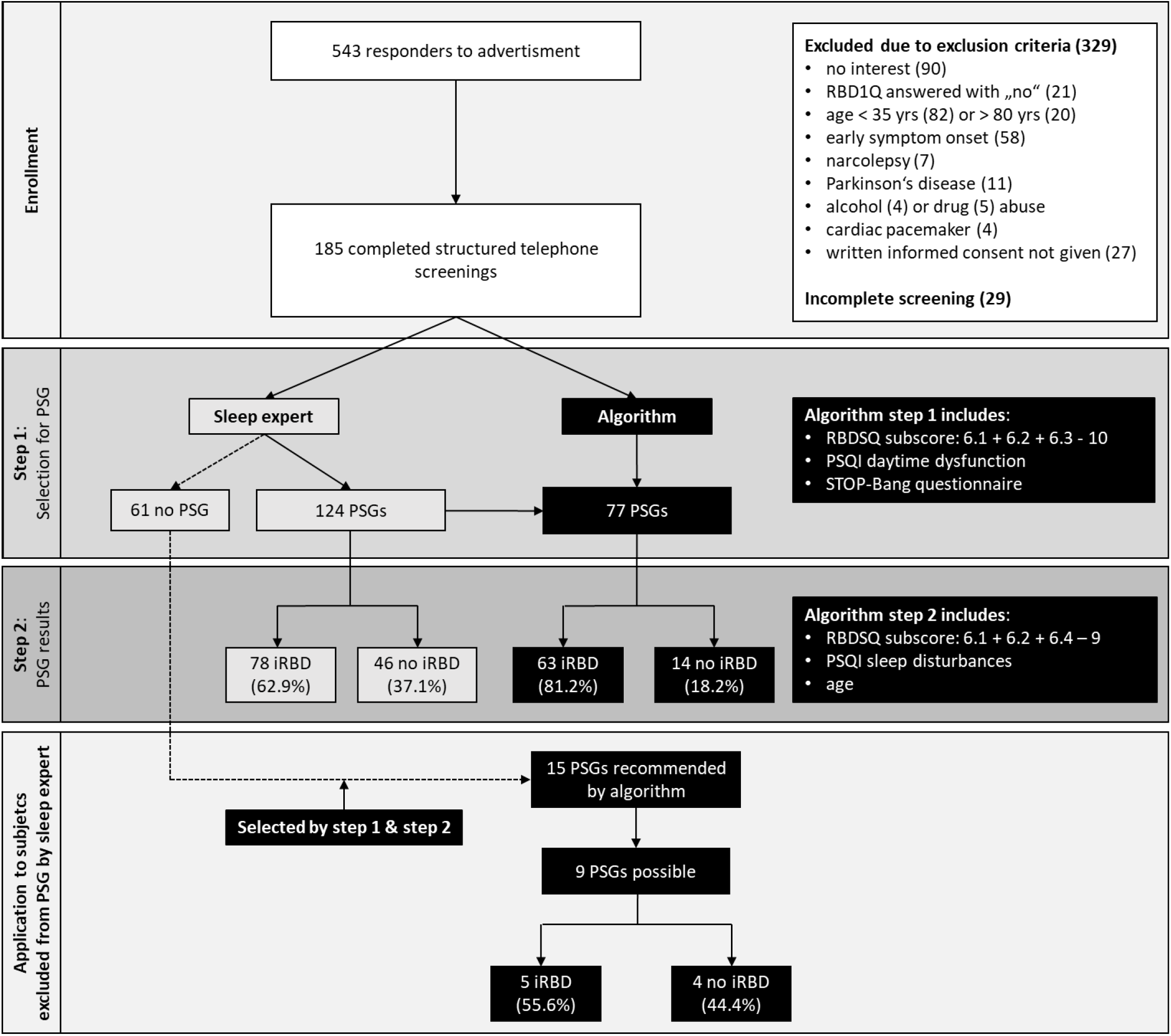
Visualization of study results. The flow chart describes the enrollment of subjects, selection steps of the sleep expert and the algorithm, and performance of the algorithm in subjects not selected for polysomnography by the sleep expert. Abbreviations: iRBD, isolated REM Sleep Behavior Disorder; PSG, polysomnography; PSQI, Pittsburgh Sleep Quality Index; RBD1Q, REM Sleep Behavior Disorder Single-Question Screen; RBDSQ, REM Sleep Behavior Disorder Screening Questionnaire; yrs, years.

### Composition of the algorithm to identify iRBD subjects

At first, we examined the performance of the RBDSQ alone as a commonly used screening tool in comparison to the expert ratings. The RBDSQ total score was a significant predictor of being chosen for PSG by the sleep expert (*p* < 0.001): considering the published RBDSQ cut-off score of >5,^8,23^ the RBDSQ showed 95.2% sensitivity and 26.2% specificity, and an AUC of 0.68. Among subjects receiving PSG, the RBDSQ total score was a significant predictor of presence of iRBD in PSG as well (*p* = 0.009), showing 96.2% sensitivity and 6.5% specificity at a cut-off score of >5 points, and an AUC of 0.64.

As the RBDSQ total score had only low specificity, we aimed to improve classification accuracy by selecting the most specific items of the RBDSQ and including information from the additionally applied questionnaires:

In the stepwise multiple regression analysis including the pre-selected items from simple logistic regression analyses for step 1 (selection of subjects to PSG after screening), the final model consisted of a RBDSQ subscore of items 6.1 + 6.2 + 6.3 – (minus) 10, the PSQI component score daytime dysfunction, and the STOP-Bang questionnaire. The final step 1 model reached 81.1% classification accuracy (sensitivity 91.9%, specificity 59%; AUC: 0.84, *p* < 0.001). The AUC of this final model significantly differed from the AUC using the RBDSQ total score only, indicating a significantly better classification accuracy of the resulting step 1 algorithm (Z = -3.3, *p* < 0.001).

In step 2 (identification of iRBD subjects upon PSG), the final model consists of the RBDSQ subscore based on items 6.1 + 6.2 + 6.4 – (minus) 9 (notably, partly different items than in step 1), age, and the PSQI component score sleep disturbances. With this 2^nd^ step 76.6% of participants were correctly classified as “iRBD” or “no iRBD” (sensitivity 83.3%, specificity 65.2%, AUC: 0.82, *p* < 0.001). The AUC of this final step 2 model significantly differed from the AUC using the RBDSQ total score alone (Z = -2.6, *p* = 0.009). Significant predictors in simple and multiple logistic regression analyses at both steps of the analysis are presented in table 2.

**Table 2.** Significant predictors of being selected for PSG (step 1) and PSG-proven iRBD (step 2) in simple and multiple logistic regression analyses.

| Characteristic | Simple regression analysis |  |  | Multiple regression analysis |  |  | Nagelkerke's R<sup>2</sup> |
| --- | --- | --- | --- | --- | --- | --- | --- |
|  | Beta Coefficient | OR (95% CI) | *P* | Beta Coefficient | OR (95% CI) | *P* |  |
| **Step 1 (chosen for PSG)** |  |  |  |  |  |  |  |
| RBDSQ Subscore (1) <sup>a</sup> | 1.23 | 3.42 (2.23–5.12) | <0.001 | 1.19 | 3.29 (2.12–5.11) | <0.001 | 0.41 |
| PSQI Daytime dysfunction | -0.78 | 0.46 (0.31–0.69) | <0.001 | -0.67 | 0.52 (0.32–0.82) | 0.005 |  |
| STOP-Bang Subscore <sup>b</sup> | -0.43 | 0.65 (0.52–0.82) | <0.001 | -0.46 | 0.63 (0.48–0.83) | 0.001 |  |
| RIS Subscore <sup>c</sup> | -0.11 | 0.90 (0.85–0.95) | <0.001 |  |  |  |  |
| RLS total | -0.18 | 0.84 (0.74–0.95) | 0.006 |  |  |  |  |
| PSQI Sleep disturbances | -0.64 | 0.53 (0.30–0.94) | 0.030 |  |  |  |  |
| **Step 2 (iRBD in PSG)** |  |  |  |  |  |  |  |
| RBDSQ Subscore (2) <sup>d</sup> | 1.26 | 3.52 (2.05–6.05) | <0.001 | 1.25 | 3.50 (1.98–6.20) | <0.001 | 0.38 |
| Age | 0.07 | 1.07 (1.03–1.12) | 0.002 | 0.07 | 1.07 (1.02–1.13) | 0.007 |  |
| PSQI Sleep disturbances | -0.91 | 0.40 (0.19–0.85) | 0.018 | -0.83 | 0.44 (0.19–1.02) | 0.055 |  |
| PSQI Sleep Quality | -1.00 | 0.34 (0.21–0.66) | <0.001 |  |  |  |  |
| RLS total | -0.19 | 0.83 (0.70–0.98) | 0.026 |  |  |  |  |
| PSQI Sleep duration | -0.74 | 0.45 (0.30–0.77) | 0.002 |  |  |  |  |
| PSQI Habitual sleep efficiency | -0.37 | 0.69 (0.48–0.99) | 0.046 |  |  |  |  |
| RIS Subscore <sup>c</sup> | -0.10 | 0.91 (0.84–0.98) | 0.017 |  |  |  |  |

In the first step, all patients who received a telephone screening were included, and ‘performed PSG’ vs. ‘no PSG’ was used as the criterion variable. In the second step, we only included patients who received PSG, and presence of iRBD confirmed by PSG was used as the criterion variable (‘iRBD’ vs. ‘no iRBD’).
<sup>a</sup> RBDSQ subscore Step 1= RBDSQ items 6.1 + 6.2 + 6.3 – (minus) 10. <sup>b</sup> STOP-Bang subscore = STOP-Bang items 1 to 7. <sup>c</sup> RIS subscore = RIS items 3 to 10. <sup>d</sup> RBDSQ subscore Step 2 = RBDSQ items 6.1 + 6.2 + 6.4 – (minus) 9
Abbreviations: ESS, Epworth Sleepiness Scale; iRBD, isolated REM Sleep Behavior Disorder; OR, Odds ratio; PSQI, Pittsburgh Sleep Quality Index; PSG, Polysomnography; RBDSQ, REM Sleep Behavior disorder Screening Questionnaire; RIS, Regensburg Insomnia Scale; RLS, Restless Legs Syndrom screening questionnaire; STOP-Bang, screening tool for obstructive sleep apnea.

To make the algorithm derived by combining the final logistic regression models from step 1 and step 2 available for general use, we created a calculator to compute the individual prediction of iRBD based on the previous named questionnaires (see supplement).

### Application of the algorithm

Following the selection criteria of our algorithm (that means combining step 1 and step 2 of the model) on all patients that underwent PSG evaluation, 77 PSGs would have been performed and 63 iRBD subjects would have been detected (Figure 1). Accordingly, comparing our algorithm to the expert selection, 77 instead of 124 PSG (62.1%) would have been carried out and 63 instead of 78 iRBD patients (80.8%) would have been identified. Hence, 32 of 46 (69.6%) unnecessary PSGs could have been avoided (Figure 2). Compared to a selection solely based on the RBDSQ cut-off >5, our combined algorithm had a significantly better accuracy of 78.1% (85.1% sensitivity and 65% specificity) and AUC of 0.83 (Z = -2.98, *p* = 0.003).

**Figure 2.**
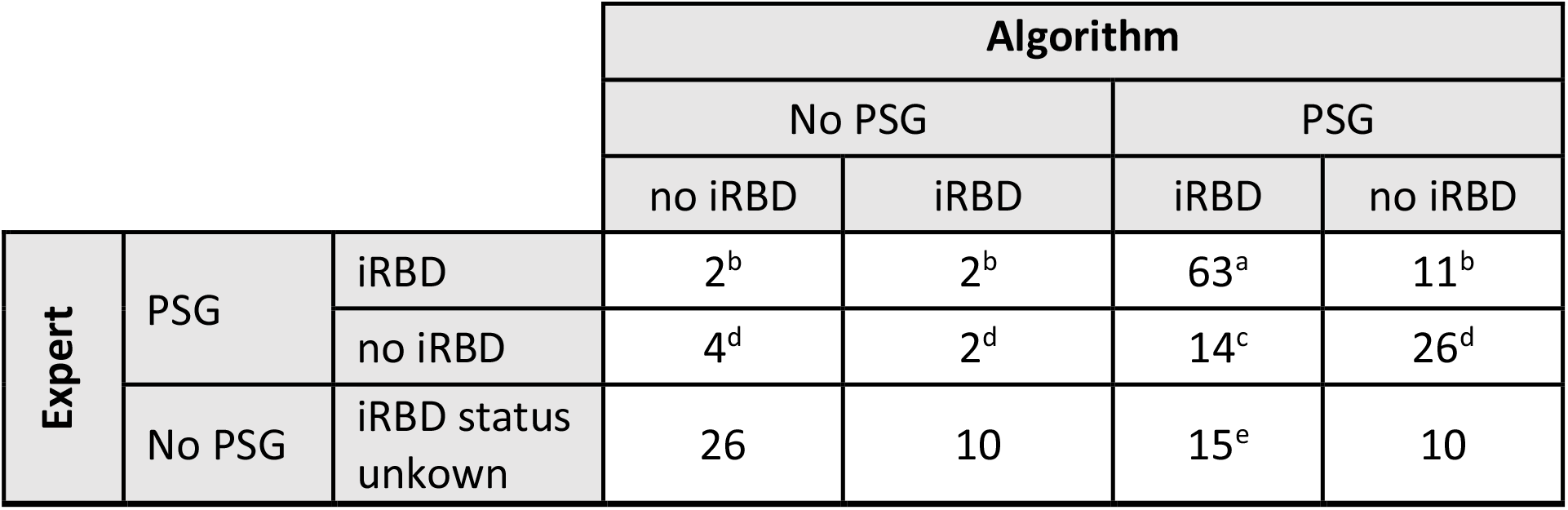
Application of the algorithm: A total of 185 subjects were evaluated by the sleep expert and the algorithm. The sleep expert could judge to select a subject for PSG (upper two rows, n = 124) or against PSG (lower row, n = 61). The algorithm judged in a similar way: first, it selected subjects against PSG examination (left columns, n = 46) or for PSG (right columns, n = 139; however, only 114 of these subjects received PSG due to sleep expert selection in the study). In a second, independent step, the algorithm evaluated potential outcome of PSG. As both steps are calculated independently, contradictory combinations (i.e., no PSG + iRBD and PSG + no iRBD) could arise. Note that the RBD-status of the subjects not allocated to PSG by the sleep-expert remains unknown (lowest row). Due to the algorithm 63^a^ subjects should have received a PSG and were correctly assigned to ‘iRBD’. In total, 15^b^ iRBD patients would have been missed using the algorithm, either they were assigned to no PSG in the 1st step, to no iRBD in the 2nd step or both. 14^c^ unnecessary PSGs would have been performed due to the algorithm. Taken together 77^a,c^ instead of 124^a-d^ PSGs (62.1%) would have been carried out and 63^a^ instead of 78^a,b^ iRBD patients (80.8%) would have been identified. Hence, 32^d^ of 46^c,d^ (69.6%) unnecessary PSGs could have been avoided. In additional 15^e^ subjects, who were excluded from PSG after the expert rating, the two-step algorithm predicted iRBD. These subjects were hereafter invited to PSG.

We applied the results of our proposed algorithm on all subjects who completed screening (n = 185). Hereby, in 15 subjects, who were excluded from PSG according to the expert rating, the two-step algorithm predicted iRBD. We subsequently contacted and invited these subjects to PSG. Nine subjects were still interested and underwent PSG. Of these, 5 subjects (56%) were diagnosed with iRBD, which were missed by the sleep expert ratings.

## Discussion

In this study, we present a novel screening algorithm to optimize identification of subjects with iRBD from the general population. Our approach does not require a selection step for PSGs by a sleep expert. The algorithm allowed to identify 80% of iRBD patients who were identified by a sleep expert at a 40% reduction in PSG numbers. Therefore, we included multiple validated sleep questionnaires and screening tools covering common aspects for differential diagnosis of iRBD within a structured screening assessment and identified the most relevant items for predicting iRBD. We were able to improve classification accuracy compared to the RBDSQ alone and facilitate a more selective use of PSGs compared to a sleep expert.

For many years, slowing down the process of neurodegeneration has been one of the main important goals in PD research. However, various clinical trials of potential neuroprotective therapies have been unsuccessful.^24–26^ One reason might be that patients with established motor PD were included in those studies. In a stage where moderate motor symptoms of PD are present, large parts of the dopaminergic neurons and their terminals are already irreversibly degenerated.^27,28^ iRBD represents an early stage of an α-synucleinopathy, hence dopaminergic and cerebral neurodegeneration has not yet progressed as far as in overt PD patients.^29,30^ It is proposed that starting a pharmacological intervention as early as possible during a neurodegenerative process will lead to the greatest benefit.^31^ iRBD is by far the most specific indicator of prodromal PD.^32,33^ Therefore, iRBD cohorts will represent the main target population for future clinical trials on potential neuroprotective strategies in PD. Thus, it appears that the identification of subjects with iRBD from the general population will become increasingly important in the near future.

Several RBD screening instruments have been developed in the past,^8,34,35^ however, they exhibit some limitations. One very common and widely used screening tool for detecting RBD is the RBDSQ. Yet, recent studies questioned the validity of the RBDSQ to assess RBD in the general population: For instance, only a low agreement between two assessments was found in a two year follow-up study aiming to evaluate the consistence of “probable RBD” diagnosis.^36^ Another study demonstrated that the diagnostic value of the RBDSQ depends on the clinical settings and the given knowledge of the respondent about RBD as a disease.^9^ Most importantly, differential diagnosis to RBD, e.g., obstructive sleep apnea (OSAS), insomnias, and restless legs syndrome are not covered by the RBDSQ. This limitation leads to a high sensitivity at the expense of a lower specificity, which is in line with its designation as a screening instrument optimized to sensitivity.^8^ The Mayo sleep questionnaire (MSQ), another RBD screening questionnaire, does cover some aspects of differential diagnosis, yet it requires information from the subject’s bed partner to exhibit high accuracy, which might not always be available.^35^ Our proposed algorithm does rely on self-reported information only similarly to the RBDSQ; however, the addition of relevant items from other questionnaires optimized accuracy. Interestingly, the RBDSQ showed a very low specificity in our analysis, which might also be due to pre-selection of subjects with the RBD1Q in our set-up.

Our results demonstrate that especially OSAS and insomnia are important differential diagnoses to consider when screening for iRBD. Wallowing around in bed, typically in insomnia, and waking up from apnea can easily be mistaken as agitated movements and eventually, mimic dream-enacting behavior as it occurs in iRBD. Furthermore, subjects with iRBD often report a good sleep quality and no significant impairment of their daytime function. In contrast subjects with insomnia often emphasize a bad sleep quality affecting their daily activities, indicated by significantly better PSQI and RIS sum scores of the patients with iRBD compared to subjects without iRBD in our screening assessment (table 1). Notably, we observed no difference regarding daytime sleepiness between all groups, as one would expect a higher extent of daytime sleepiness in iRBD as an early symptom of α-synucleinopathy.^32,33^

It must be pointed out that there are controversies regarding screening for iRBD and whether iRBD patients should be informed about their risk of phenoconversion.^37^ Given the fact that no disease modifying therapies or preventive strategies exist so far, the diagnosis of iRBD might lead to anxiety and hopelesseness.^38,39^ Furthermore, it is difficult to determine if and when motor symptoms appear, leaving patients with continuing uncertainty about their future health. On the other hand, it is argued that patients have the right to know about their clinical condition and their potential risk of developing a neurodegenerative disorder which can then be taken into consideration regarding future life plans.^39^ Also, subjects with iRBD in research cohorts are usually offered a close medical attendance and they are provided an early treatment of symptoms when they occur, which could impact quality of life.^40^ It is suggested that disclosure of iRBD diagnosis and the concomitant risks should be individually based on the patients wish to be informed and possible comorbidities, e.g., psychiatric disorders.^39,41^ The expert opinions are in accordance with the results of a recently published study evaluating patients with PD about their opinion.^42^ Initially PD patients seemed to be skeptical, in particular due to the absence of modifying therapies. However, most of the patients would agree to know about iRBD diagnosis and its risks under certain circumstances, namely recommendations on lifestyle changes, an early assessment of the patients wish to know and regular medical attendance.^42^ In our study, we permitted early drop-out after education about the study aims and potential iRBD diagnosis before any screening and PSG examination started, which is line with recently published recommendations.^41^ Our study has some limitations. First, we only used the RBDSQ as a specific RBD questionnaire and did not include further questionnaires for comparison, e.g., the MSQ or the Innsbruck RBD inventory.^34,35^ However, accuracy of the MSQ is strongly dependent of information from bed-partners,^36^ which we by purpose did not want to include. Additionally, the results of the analysis of the RBDSQ have to be considered with caution as the RBDSQ was applied on an already pre-selected cohort after education about study purpose including the background of iRBD.^9^ The first step of our two-step algorithm is based on experiences and subjective evaluation of our sleep expert, and we might have missed some iRBD subjects by this selection. So far, we could only perform a limited validation of our algorithm on a small amount of people (n=9) as we had no validation cohort. We will use the algorithm in our ongoing enlargement of our local cohort for future validation.

Still, our proposed algorithm is the first solely self-reported questionnaire-based assessment tested to identify iRBD patients from the general population and showed superior accuracy to the commonly used RBDSQ. We could demonstrate that a substantial number of PSG examinations could have been saved by using the algorithm without relevant decline in detection of iRBD subjects, which eventually reduce costs.

In summary, our model provides a convenient tool for research studies and clinical application to identify subjects with iRBD from the public. Such a tool allows for cost-efficient screening of larger groups for potentially upcoming disease-modifying trials.

## Supporting information

Supplemental Algorithm

## Data Availability

All data produced in the present study are available upon reasonable request to the authors.

## Acknowledgment

We thank all study participants for their participation. Furthermore, we appreciate the great support of Nora Pagel and Nora Sembowski.

## Authors’ Roles

1. Research project: A. Conception, B. Organization, C. Execution;
2. Statistical Analysis: A. Design, B. Execution, C. Review and Critique;
3. Manuscript: A. Writing of the first draft, B. Review and Critique.

**AS:** 1B, 1C, 2A, 2B, 3A; **AO**: 2A, 2B, 3B; **WH:** 1B, 1C, 3B; **CEJD:** 1A, 1B, 1C, 2C, 3B; **ML:** 1B, 1C, 3B; **CB:** 1B, 1C, 3B; **JK:** 1B, 1C, 3B; **HSD**: 3B; **WHO:** 3B; **GRF**: 1A, 3B; **STJ:** 2A, 2B, 3A **MS:** 1A, 1B, 1C, 2A, 2B, 3A

