## Supplemental Table 1 for "Evaluation of a structured screening assessment to detect patients with isolated REM Sleep Behavior Disorder"

### Supplementary Material

#### Supplementary Figure 1:

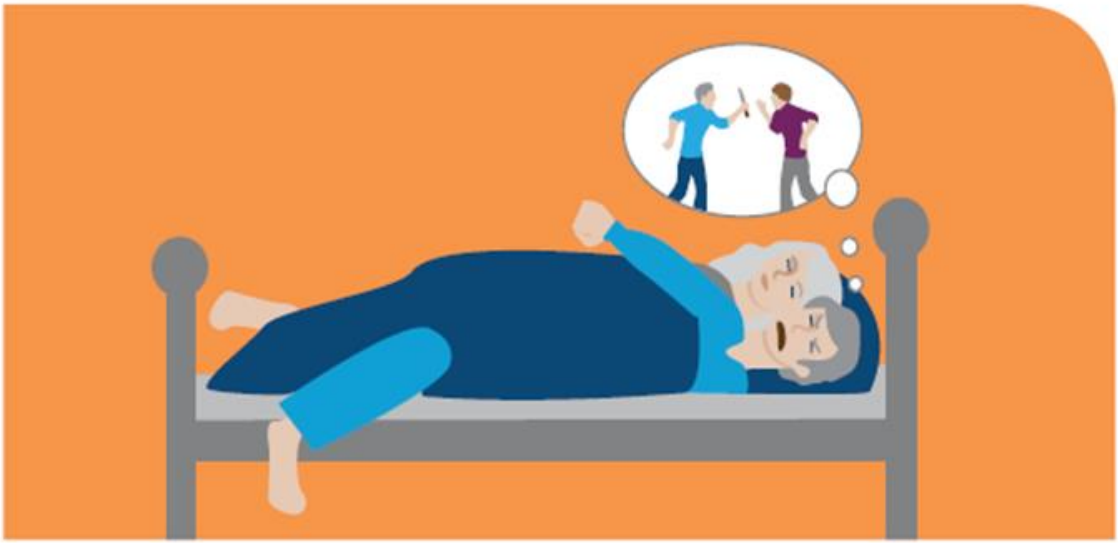

**Graphical advertisement for recruiting subjects with iRBD placed in local newspaper: Elderly man with dream-enacting behavior and violent dream content**. Additionally, the German version of the single-question screen for RBD (RBD1Q; "Did you notice or did someone else notice that you do act out your dreams?") was added underneath the graphic. Subjects who could affirm to the RBD1Q were encouraged to contact our research group.
